# Impacts of the COVID-19 pandemic on sepsis incidence, etiology and hospitalization costs in France: a retrospective observational study

**DOI:** 10.1101/2024.09.11.24311990

**Authors:** Marie Al Rahmoun, Alexandre Sabaté-Elabbadi, Didier Guillemot, Christian Brun-Buisson, Laurence Watier

## Abstract

**Background:** Sepsis is a serious medical condition that causes long-term morbidity and high mortality, annually affecting millions of people worldwide. The COVID-19 pandemic may have impacted its burden.

**Objective:** To estimate the impact of the COVID-19 pandemic on sepsis incidence, etiology and associated hospitalization costs in metropolitan France.

**Methods:** This retrospective observational study used data drawn from a cohort of hospitalized sepsis patients in France’s national healthcare database. Sepsis was identified through both explicit ICD-10 codes (E-sepsis) and implicit codes (I-sepsis). Participants included all patients aged 15 years or older hospitalized with E-sepsis or I-sepsis in metropolitan France between January 1, 2018, and December 31, 2022. Patient and hospital stay characteristics were described by sepsis type (E-sepsis, I-sepsis) and overall. The distribution of sepsis etiology was estimated for each year. Annual incidence rates were estimated overall and by sepsis type and etiology. Total and median per-stay hospitalization costs were calculated.

**Results:** The total age- and sex-standardized sepsis incidence rate per 100,000 increased slightly from 2018 (446, 95% CI 444.2 to 447.7) to 2020 (457, 95% CI 455.1 to 458.6) and then decreased in 2022 (382, 95% CI 380.2-383.7) (p <.0001). Incidence rates decreased for both E-sepsis and bacterial sepsis during the pandemic period, whereas I-sepsis incidence increased in 2020 and 2021, associated with a marked increase in viral sepsis and co-infections (p <.0001 for E- and I-sepsis). Viral sepsis represented about 10% of all sepsis cases during the pandemic, but only about 1% prior to the pandemic. Total sepsis-associated hospitalization costs and extra medication costs increased during the pandemic. Characteristics of patients and their hospital stays were overall stable over the five-year study period.

**Conclusions:** The COVID-19 pandemic led to a higher burden of sepsis in French hospitals and an increase in hospital stay costs. Critically, our study highlights the need for introducing explicit viral sepsis codes within the ICD-11 classification system and for achieving a consensus on its definition in order to robustly estimate sepsis incidence.

## Introduction

Sepsis is a complex condition recognized as a major global public health problem due to its long-term morbidity and high mortality ^1,2^. It also incurs a high economic burden for healthcare systems in many countries ^3,4^. According to the Third International Consensus Definitions of Sepsis and Septic Shock (Sepsis-3), sepsis is a dysregulated host response to infection, whether bacterial, viral, fungal or parasitic, resulting in life-threatening organ dysfunction ^5^. However, several changes in diagnosis, documentation and coding practices for sepsis and organ dysfunction have occurred over the years ^6^. Although many clinicians associate sepsis with severe bacterial infection, severe viral infection – including severe acute respiratory syndrome coronavirus 2 (SARS-CoV-2) infection causing coronavirus disease 2019 (COVID-19) – often manifests as respiratory dysfunction and can directly or indirectly impair other organs ^7–9^. This is consistent with viral sepsis according to Sepsis-3 criteria ^10,11^.

In France, a nationwide, population-based, retrospective observational study showed that the annual incidence of bacterial sepsis between 2015 and 2019 was around 350-400 cases per 100 000 inhabitants ^12^. In this study, the proportion of sepsis of viral or fungal etiology, without bacterial coinfection, was estimated at only 1.7% of all sepsis cases.

The COVID-19 pandemic has likely modified the distribution of sepsis etiology, since a substantial proportion of hospitalized COVID-19 cases are associated with sepsis ^13–15^. There is a lack of data regarding the epidemiology of virus-associated sepsis, including COVID-19-associated sepsis, because of the common misperception that sepsis can only be caused by bacterial pathogens. This is reflected by the absence of virus-associated sepsis codes in both the tenth and eleventh revisions of the International Classification of Diseases (ICD-10 and 11) diagnostic coding system ^16^. However, a meta-analysis of 57 studies found that most COVID-19 patients admitted to the intensive care unit (ICU) met Sepsis-3 criteria, despite a high heterogeneity of sepsis prevalence across included studies ^17^.

In this study, we aimed to evaluate the impact of the COVID-19 pandemic on sepsis incidence, etiology and associated hospitalization costs in metropolitan France. We used a national hospitalized patient database to estimate the distribution of different infectious agents (bacteria, viruses, fungi and parasites) associated with sepsis and describe the characteristics of sepsis patients and their hospital stays before and during the COVID-19 pandemic.

## Materials and Methods

### Data source

This is a retrospective, observational study of hospitalized patients with sepsis, regardless of etiology, registered in the French National Hospital Discharge Database (Programme de Médicalisation des Systèmes d’Information, PMSI) issued from the French healthcare database (Système National des Données de Santé, SNDS) (**eMethods**). These data cover all acute care hospitalizations in French public and private hospitals. Hospitalization costs and COVID-19 vaccination data were also obtained from this same database. Demographic data for incidence rates estimations were obtained from the French Census of the National Institute of Statistics and Economic Studies ^18^.

### Study population

The study included all patients aged 15 years or older hospitalized with sepsis between 1 January 2018 and 31 December 2022 in metropolitan France (excluding overseas French territories). Stays of less than 1 day and discharged alive were excluded. When multiple stays per year for a patient were observed, only the last one was included to avoid underestimating in-hospital mortality. In this article, when mentioned, the pre-pandemic period refers to 2018-2019 while the pandemic period refers to 2020-2022.

Sepsis was defined as either of two mutually exclusive types of sepsis, explicit sepsis (hereafter E-sepsis) or implicit sepsis (I-sepsis), similar to definitions used in previous studies on the same French healthcare database and others ^1,12,19^, with some changes to meet new coding recommendations of the Technical Agency for Information on Hospital Care in 2021 (ATIH) ^20^ (**eMethods)**.

E-sepsis was defined as a hospital stay with one of the selected ICD-10 codes for sepsis as primary diagnosis (PD: condition requiring hospitalization), related diagnosis (RD: adds information to PD) or significant associated diagnosis (SAD: complications and comorbidities potentially affecting the course or cost of hospitalization). I-sepsis was defined as a hospital stay with one of the selected ICD-10 codes for infection (other than those defining E-sepsis) as PD, RD or SAD with two associated conditions: intensive care unit (ICU) admission and at least one of the selected ICD-10 codes for organ dysfunction or one or more codes for organ support from the Common Classification of Medical Acts (CCAM) (**eTable 1**). Etiology of sepsis was classified by ICD-10 codes (**eTable 1**). COVID-19 hospitalizations, which fall into the definition of I-sepsis patients when admitted to ICU and having organ dysfunction/support, were defined as a hospital stay with one of the ICD-10 codes for COVID-19.

### Covariates

The socio-demographic and clinical characteristics of patients and their hospital stay characteristics were extracted from the data, including sex, age, Charlson index, detailed comorbidities, infection sites, admission source, septic shock, ICU admission, length of stay, and hospital discharge. Using the ICD-10 codes list defined by Pandolfi et al ^12^, a total of 14 infection sites were identified: lower respiratory tract, blood, urinary and genital tracts, gastrointestinal and abdomen, heart and mediastinum, skin and soft tissues, medical devices, bones and joints, nervous system, ears, nose and throat, pregnancy, eyes, multiple sites, and unknown.

### Statistical analysis

The overall annual incidence rate, expressed as the number of cases per 100,000 inhabitants relative to the general population (crude and sex and age-adjusted based on 2022 population distribution), the incidence rate for each sex and age category, and their 95% confidence intervals (CIs) were estimated.

The distributions of presumed causal infectious agents of sepsis (bacteria, viruses, fungi or parasites, and co-infections) were estimated in each year. Based on these estimates, etiology-specific sepsis incidence rates were also estimated.

The total and median (IQR) per-stay direct hospital costs from the provider perspective were calculated, per sepsis type and overall. Supplementary costs for expensive medications not included in the cost of the hospital stay, like medications subject to temporary authorization for use and thrombolytic drugs for the treatment of ischemic stroke, were also calculated as extra medication costs.

Patient and hospital stay characteristics were described by sepsis type (E-sepsis or I-sepsis) and overall by year. The percentage of cases with admission to ICU and septic shock were described only for E-sepsis, as admission to ICU was part of the selection criteria for I-sepsis while septic shock code was only part of the E-sepsis definition. Number and percentage were reported for qualitative variables, and median and interquartile range (IQR) for quantitative variables. No confidence intervals (CI) were calculated for percentages as the data cover the national population. The COVID-19 vaccination coverage rate of the study population was also estimated for 2021 and 2022.

A Cochran-Armitage test for trend was used to assess change in incidence rate and in-hospital mortality over time. Analyses were performed using SAS Enterprise Guide 8.

## Results

### Type of sepsis

The majority of sepsis patients (around 90%) had only one same-year hospital stay related to sepsis, and less than 2.5% had more than 2 sepsis-related stays in the same year (**eTable 2**). Before the COVID-19 pandemic, the annual number of sepsis patients in metropolitan France was stable at around 230,000 (**Table 1**). This number increased slightly in the first two years of the pandemic (242,175 cases in 2020 and 236,076 in 2021) and then decreased markedly in 2022 (207,409 cases). This increase in sepsis cases during the pandemic was due to increasing I-sepsis cases, from 58,293 in 2019 to 82,292 in 2021, whereas E-sepsis decreased continuously from 2019 to 2022.

**Table 1.** Characteristics of patients with sepsis according to sepsis type (E-sepsis/I-sepsis)^a^ in metropolitan France, 2018-2022.

|  |  | 2018 |  | 2019 |  | 2020 |  | 2021 |  | 2022 |  |
| --- | --- | --- | --- | --- | --- | --- | --- | --- | --- | --- | --- |
|  |  | (n=229,668) |  | (n=232,355) |  | (n=242,175) |  | (n=236,076) |  | (n=207,409) |  |
|  |  | E-sepsis | I-sepsis | E-sepsis | I-sepsis | E-sepsis | I-sepsis | E-sepsis | I-sepsis | E-sepsis | I-sepsis |
|  |  | (n=170,480) | (n=59,188) | (n=174,062) | (n=58,293) | (n=168,953) | (n=73,222) | (n=153,784) | (n=82,292) | (n=144,036) | (n=63,373) |
| **Patient's characteristics, n (%)** |  |  |  |  |  |  |  |  |  |  |  |
| **Sex** |  |  |  |  |  |  |  |  |  |  |  |
|  | Male | 97,645 (57.3) | 34,989 (59.1) | 99,811 (57.3) | 34,582 (59.3) | 99,375 (58.8) | 46,112 (63.0) | 91,211 (59.3) | 51,522 (62.6) | 84,700 (58.8) | 39,252 (61.9) |
|  | Female | 72,835 (42.7) | 24,199 (40.9) | 74,251 (42.7) | 23,711 (40.7) | 69,578 (41.2) | 27,110 (37.0) | 62,573 (40.7) | 30,770 (37.4) | 59,336 (41.2) | 24,121 (38.1) |
| **Age (years)** |  |  |  |  |  |  |  |  |  |  |  |
|  | 15-29 | 4,845 (2.8) | 1,888 (3.2) | 4,807 (2.8) | 1,853 (3.2) | 4,220 (2.5) | 1,845 (2.5) | 3,710 (2.4) | 2,315 (2.8) | 3,617 (2.5) | 2,045 (3.2) |
|  | 30-44 | 8,853 (5.2) | 3,802 (6.4) | 8,777 (5.0) | 3,528 (6.1) | 8,077 (4.8) | 4,615 (6.3) | 7,494 (4.9) | 6,466 (7.9) | 6,917 (4.8) | 4,184 (6.6) |
|  | 45-54 | 12,919 (7.6) | 5,697 (9.6) | 12,989 (7.5) | 5,414 (9.3) | 12,174 (7.2) | 7,841 (10.7) | 11,028 (7.2) | 9,995 (12.2) | 9,506 (6.6) | 6,206 (9.8) |
|  | 55-64 | 26,623 (15.6) | 10,782 (18.2) | 26,373 (15.2) | 10,590 (18.2) | 25,914 (15.3) | 14,360 (19.6) | 23,321 (15.2) | 17,574 (21.4) | 20,868 (14.5) | 12,074 (19.1) |
|  | 65-74 | 41,363 (24.3) | 15,468 (26.1) | 42,958 (24.7) | 15,695 (26.9) | 43,889 (26.0) | 20,957 (28.6) | 40,956 (26.6) | 24,319 (29.6) | 37,166 (25.8) | 18,780 (29.6) |
|  | 75-84 | 41,466 (24.3) | 13,703 (23.2) | 42,139 (24.2) | 13,332 (22.9) | 40,902 (24.2) | 15,855 (21.7) | 36,801 (23.9) | 15,429 (18.8) | 36,022 (25.0) | 13,841 (21.8) |
|  | ≥ 85 | 34,411 (20.2) | 7,848 (13.3) | 36,019 (20.7) | 7,881 (13.5) | 33,777 (20.0) | 7,749 (10.6) | 30,474 (19.8) | 6,194 (7.5) | 29,940 (20.8) | 6,243 (9.9) |
|  | Median [IQR] | 72 [61-83] | 69 [58-79] | 72 [62-83] | 70 [59-79] | 72 [62-83] | 69 [58-77] | 72 [62-82] | 67 [56-75] | 73 [63-83] | 69 [58-77] |
| **Charlson index** |  |  |  |  |  |  |  |  |  |  |  |
|  | 0 | 59,407 (34.9) | 18,609 (31.4) | 60,919 (35.0) | 18,150 (31.1) | 58,614 (34.7) | 29,357 (40.1) | 52,821 (34.4) | 38,570 (46.9) | 47,237 (32.8) | 21,587 (34.1) |
|  | 1-2 | 59,336 (34.8) | 22,751 (38.4) | 60,593 (34.8) | 22,701 (38.9) | 58,735 (34.8) | 26,482 (36.2) | 53,339 (34.7) | 26,892 (32.7) | 50,515 (35.1) | 24,463 (38.6) |
|  | 3-4 | 23,101 (13.6) | 11,084 (18.7) | 23,578 (13.6) | 10,899 (18.7) | 23,695 (14.0) | 10,874 (14.9) | 22,368 (14.6) | 10,646 (12.9) | 22,364 (15.5) | 10,837 (17.1) |
|  | ≥ 5 | 28,636 (16.8) | 6,744 (11.4) | 28,972 (16.6) | 6,543 (11.2) | 27,909 (16.5) | 6,509 (8.9) | 25,256 (16.4) | 6,184 (7.5) | 23,920 (16.6) | 6,486 (10.2) |
|  | Median [IQR] | 2 [0-3] | 2 [0-3] | 2 [0-3] | 2 [0-3] | 2 [0-3] | 1 [0-2] | 2 [0-3] | 1 [0-2] | 2 [0-3] | 2 [0-3] |

|  | 2018 |  | 2019 |  | 2020 |  | 2021 |  | 2022 |  |
| --- | --- | --- | --- | --- | --- | --- | --- | --- | --- | --- |
|  | (n=229,668) |  | (n=232,355) |  | (n=242,175) |  | (n=236,076) |  | (n=207,409) |  |
|  | E-sepsis<br>(n=170,480) | I-sepsis<br>(n=59,188) | E-sepsis<br>(n=174,062) | I-sepsis<br>(n=58,293) | E-sepsis<br>(n=168,953) | I-sepsis<br>(n=73,222) | E-sepsis<br>(n=153,784) | I-sepsis<br>(n=82,292) | E-sepsis<br>(n=144,036) | I-sepsis<br>(n=63,373) |
| **Comorbidities** |  |  |  |  |  |  |  |  |  |  |
| Cancer | 46,209 (27.1) | 9,980 (16.9) | 46,605 (26.8) | 9,697 (16.6) | 44,603 (26.4) | 9,867 (13.5) | 39,269 (25.5) | 9,915 (12.1) | 36,797 (25.6) | 10,380 (16.4) |
| Congestive heart failure | 32,209 (18.9) | 17,568 (29.7) | 32,733 (18.8) | 17,124 (29.4) | 32,743 (19.4) | 17,481 (23.9) | 32,208 (20.9) | 16,666 (20.3) | 32,341 (22.5) | 16,542 (26.1) |
| Renal disease | 22,549 (13.2) | 8,050 (13.6) | 23,501 (13.5) | 8,217 (14.1) | 23,517 (13.9) | 9,102 (12.4) | 22,747 (14.8) | 8,928 (10.9) | 22,606 (15.7) | 8,735 (13.8) |
| Chronic pulmonary disease | 15,238 (8.9) | 11,859 (20.0) | 15,548 (8.9) | 11,997 (20.6) | 15,732 (9.3) | 12,626 (17.2) | 15,087 (9.8) | 12,879 (15.7) | 14,990 (10.4) | 13,216 (20.9) |
| Metastatic carcinoma | 20,015 (11.7) | 3,363 (5.7) | 20,342 (11.7) | 3,334 (5.7) | 19,479 (11.5) | 3,371 (4.6) | 17,117 (11.1) | 3,223 (3.9) | 15,837 (11.0) | 3,277 (5.2) |
| Diabetes with chronic complications | 10,150 (6.0) | 3,622 (6.1) | 10,170 (5.8) | 3,655 (6.3) | 10,050 (6.0) | 4,170 (5.7) | 9,179 (6.0) | 4,091 (5.0) | 8,545 (5.9) | 3,380 (5.3) |
| Paraplegia or hemiplegia | 8,780 (5.2) | 4,490 (7.6) | 8,953 (5.1) | 4,358 (7.5) | 9,070 (5.4) | 4,738 (6.5) | 8,461 (5.5) | 4,882 (5.9) | 7,571 (5.3) | 4,590 (7.2) |
| Dementia | 9,935 (5.8) | 1,930 (3.3) | 9,994 (5.7) | 1,820 (3.1) | 9,339 (5.5) | 1,966 (2.7) | 8,252 (5.4) | 1,439 (1.8) | 8,181 (5.7) | 1,389 (2.2) |
| Mild liver disease | 9,467 (5.6) | 2,959 (5.0) | 9,868 (5.7) | 2,911 (5.0) | 9,860 (5.8) | 3,150 (4.3) | 9,003 (5.9) | 3,393 (4.1) | 8,748 (6.1) | 3,115 (4.9) |
| Moderate or severe liver disease | 4,610 (2.7) | 1,290 (2.2) | 4,644 (2.7) | 1,195 (2.1) | 4,503 (2.7) | 1,254 (1.7) | 4,116 (2.7) | 1,204 (1.5) | 4,210 (2.9) | 1,284 (2.0) |
| Rheumatological disease | 2,102 (1.2) | 757 (1.3) | 2,163 (1.2) | 780 (1.3) | 2,065 (1.2) | 980 (1.3) | 1,988 (1.3) | 995 (1.2) | 1,880 (1.3) | 895 (1.4) |
| AIDS | 735 (0.4) | 378 (0.6) | 690 (0.4) | 343 (0.6) | 664 (0.4) | 364 (0.5) | 529 (0.3) | 366 (0.4) | 524 (0.4) | 306 (0.5) |
| **Infection sites** |  |  |  |  |  |  |  |  |  |  |
| Multiple sites | 35,502 (20.8) | 15,577 (26.3) | 36,487 (21.0) | 15,231 (26.1) | 35,887 (21.2) | 13,933 (19.0) | 32,755 (21.3) | 13,916 (16.9) | 32,346 (22.5) | 14,414 (22.7) |
| Lower respiratory tract | 20,479 (12.0) | 25,713 (43.4) | 20,203 (11.6) | 25,524 (43.8) | 25,673 (15.2) | 45,100 (61.6) | 26,473 (17.2) | 55,693 (67.7) | 23,481 (16.3) | 34,672 (54.7) |
| Blood | 36,533 (21.4) | - | 36,634 (21.1) | - | 33,001 (19.5) | - | 29,611 (19.3) | - | 27,796 (19.3) | - |
| Urinary and genital tract | 29,197 (17.1) | 5,748 (9.7) | 30,508 (17.5) | 5,396 (9.3) | 28,812 (17.1) | 4,610 (6.3) | 25,026 (16.3) | 3,946 (4.8) | 22,896 (15.9) | 3,847 (6.1) |
| Gastrointestinal and abdomen | 11,296 (6.6) | 2,936 (5.0) | 11,571 (6.7) | 2,945 (5.1) | 11,127 (6.6) | 2,556 (3.5) | 9,858 (6.4) | 2,438 (3.0) | 8,932 (6.2) | 2,348 (3.7) |
| Heart and mediastinum | 9,515 (5.6) | 1,733 (2.9) | 9,785 (5.6) | 1,761 (3.0) | 8,947 (5.3) | 1,645 (2.3) | 7,043 (4.6) | 1,760 (2.1) | 5,978 (4.2) | 2,084 (3.3) |
| Skin and soft tissues | 10,423 (6.1) | 517 (0.9) | 10,670 (6.1) | 535 (0.9) | 9,559 (5.7) | 364 (0.5) | 8,689 (5.7) | 321 (0.4) | 8,384 (5.8) | 359 (0.6) |

|  | 2018 |  | 2019 |  | 2020 |  | 2021 |  | 2022 |  |
| --- | --- | --- | --- | --- | --- | --- | --- | --- | --- | --- |
|  | (n=229,668) |  | (n=232,355) |  | (n=242,175) |  | (n=236,076) |  | (n=207,409) |  |
|  | E-sepsis<br>(n=170,480) | I-sepsis<br>(n=59,188) | E-sepsis<br>(n=174,062) | I-sepsis<br>(n=58,293) | E-sepsis<br>(n=168,953) | I-sepsis<br>(n=73,222) | E-sepsis<br>(n=153,784) | I-sepsis<br>(n=82,292) | E-sepsis<br>(n=144,036) | I-sepsis<br>(n=63,373) |
| Medical devices | 5,244 (3.1) | 2,621 (4.4) | 5,464 (3.1) | 2,445 (4.2) | 5,054 (3.0) | 2,019 (2.8) | 4,112 (2.7) | 2,059 (2.5) | 3,680 (2.6) | 1,729 (2.7) |
| Bones and joints | 3,979 (2.3) | 737 (1.3) | 4,237 (2.4) | 768 (1.3) | 3,761 (2.2) | 640 (0.9) | 3,360 (2.2) | 681 (0.8) | 3,241 (2.3) | 684 (1.1) |
| Nervous system | 570 (0.3) | 327 (0.6) | 590 (0.3) | 341 (0.6) | 418 (0.3) | 234 (0.3) | 384 (0.3) | 221 (0.3) | 387 (0.3) | 326 (0.5) |
| Ears, nose and throat | 328 (0.2) | 36 (0.1) | 332 (0.2) | 33 (0.1) | 218 (0.1) | 26 (0.0) | 208 (0.1) | 11 (0.0) | 196 (0.1) | 27 (0.0) |
| Pregnancy | 314 (0.2) | 0 (0.0) | 313 (0.2) | 1 (0.0) | 282 (0.2) | 1 (0.0) | 268 (0.2) | 1 (0.0) | 265 (0.2) | 1 (0.0) |
| Eyes | 32 (0.0) | 2 (0.0) | 26 (0.0) | 4 (0.0) | 24 (0.0) | 0 (0.0) | 28 (0.0) | 0 (0.0) | 20 (0.0) | 3 (0.0) |
| Unknown | 7,068 (4.2) | 3,241 (5.5) | 7,242 (4.2) | 3,309 (5.7) | 6,190 (3.7) | 2,094 (2.9) | 5,969 (3.9) | 1,245 (1.5) | 6,434 (4.5) | 2,879 (4.5) |

Abbreviations: IQR, interquartile range ; ICU, intensive care unit.
<sup>a</sup> E-sepsis: Explicit sepsis, I-sepsis: Implicit sepsis.

### Sepsis incidence rates

The total sex- and age-standardized sepsis incidence rate per 100,000 slightly increased from 2018 (446, 95% CI 444.2 to 447.7) to 2020 (457, 95% CI 455.1 to 458.6) and then decreased in 2022 (382, 95% CI 380.2-383.7) (p <.0001) (**Table 2** and **Figure 1**). The incidence rate of E-sepsis decreased during the pandemic period, whereas I-sepsis incidence increased, especially in 2020 and 2021 (p <.0001 for both E- and I-sepsis). Incidence of bacterial sepsis, including those from respiratory causes, decreased during the pandemic [from 410 (95% CI, 408.5-411.9) in 2019 to 323 (95% CI, 321.4-324.4) in 2021] while there were marked increases in the share of sepsis with viral etiology [from 6.3 (95% CI, 6.1-6.5) in 2019 to 59.3 (95% CI, 58.6-59.9) in 2021] and associated with co-infection [from 13.5 (95% CI, 13.2-13.8) in 2019 to 51.0 (95% CI, 50.4-51.6%) in 2021].

**Figure 1.**
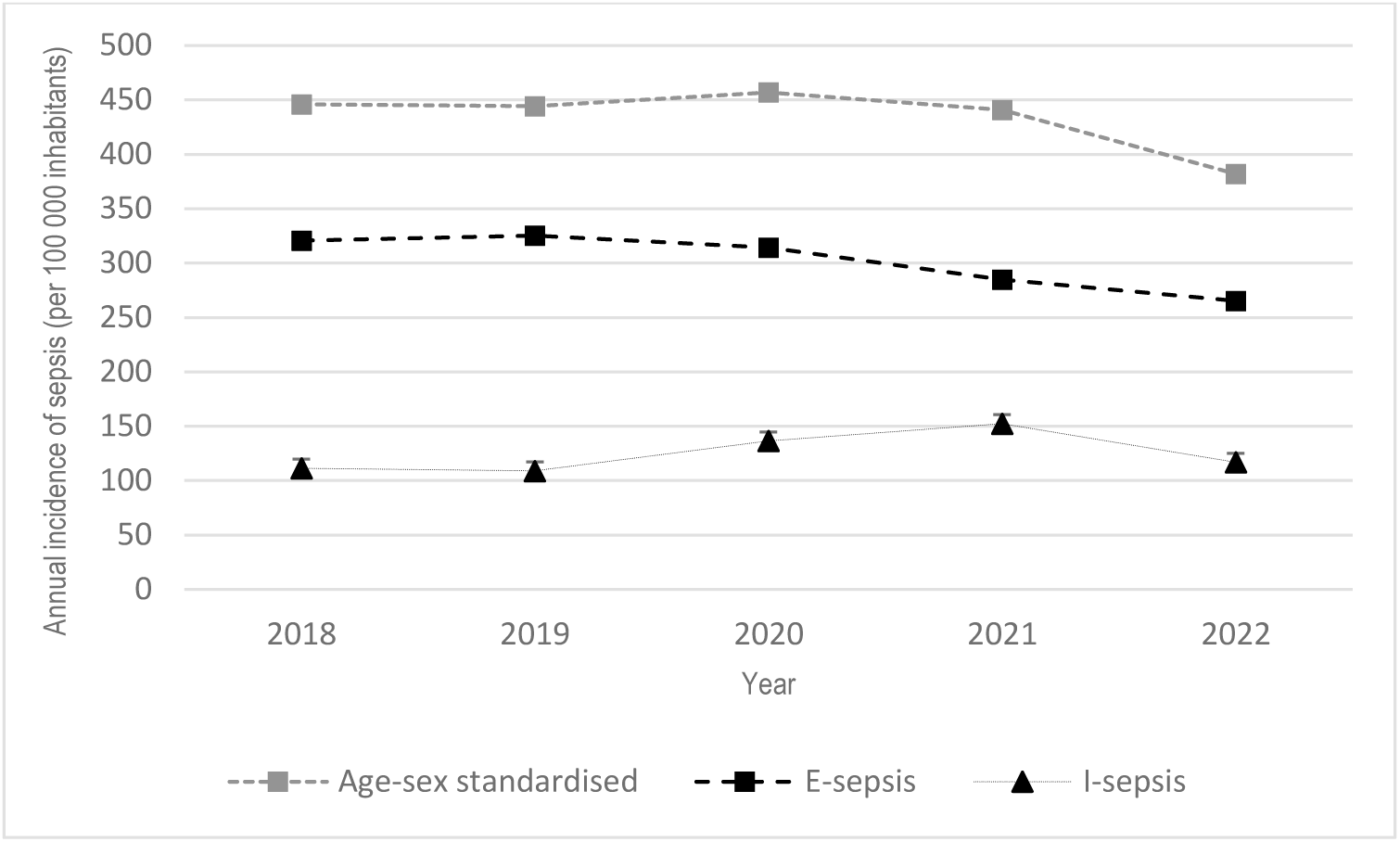
Sepsis incidence rates, overall (age-sex standardized) and by type, in metropolitan France, 2018-2022.

**Table 2.**
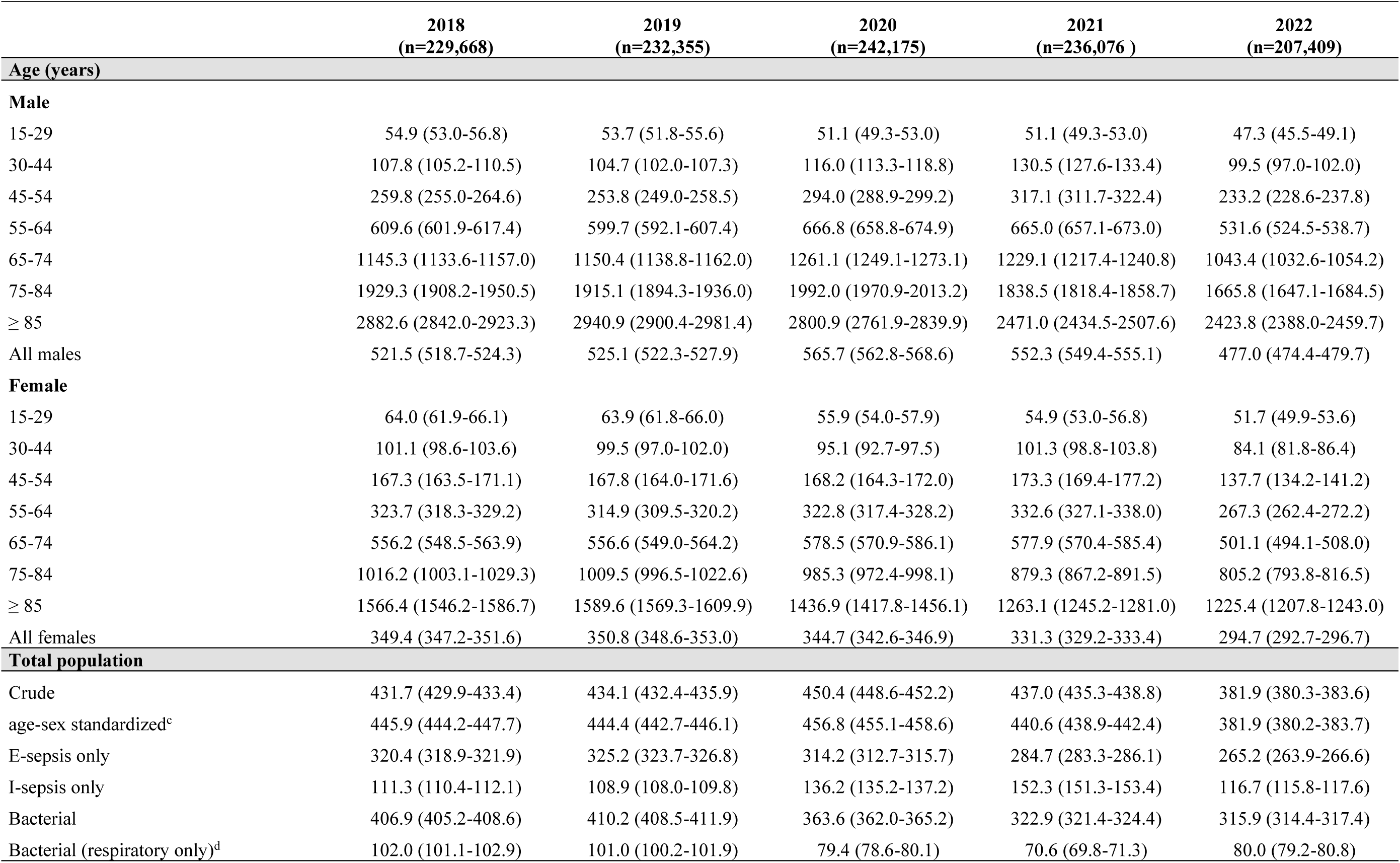
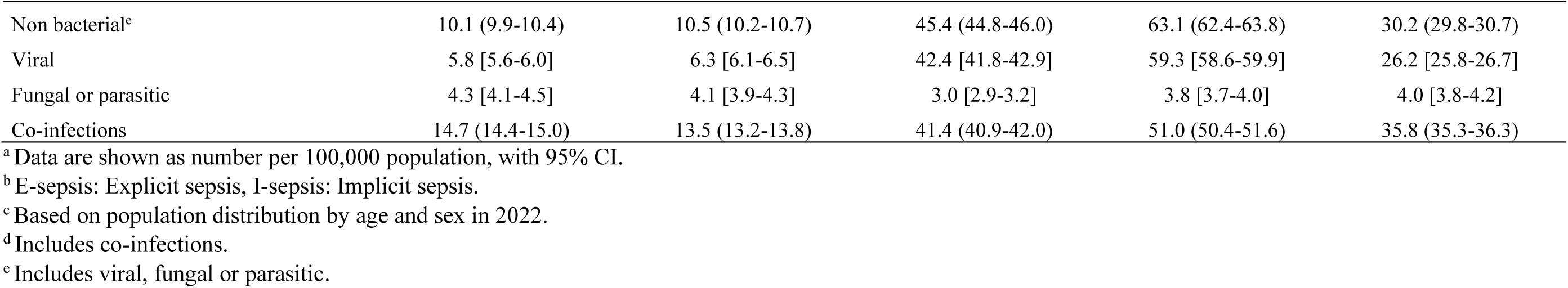
Overall sepsis incidence rates^a^ by sex and age, and according to sepsis type (E-sepsis/I-sepsis)^b^ and etiology, in metropolitan France, 2018-2022.

Throughout the study period, annual sepsis incidence remained higher for males (477, 95% CI 474.4-479.7, in 2022) than females (295, 95% CI 292.7-296.7, in 2022) and was considerably higher for people aged 55 years and older.

### Sepsis etiology

Bacterial sepsis before the COVID-19 pandemic was stable (almost 95%), decreased in 2020 and 2021 (80.7% and 73.9%, respectively) and rose in 2022 (82.7%) (**Figure 2**). Conversely, sepsis of viral etiology increased from 1.5% in 2018 and 2019 to 9.4% in 2020 and 13.6% in 2021, then decreased to 6.9% in 2022. Similar evolution was observed for co-infections. Viral sepsis relative increase during the pandemic was particularly high among I-sepsis: from around 5.5% in 2018-2019 to 31.1% in 2020 and 38.9% in 2021, before decreasing to 22.4% in 2022. Accordingly, bacterial sepsis decreased markedly among I-sepsis while remaining stable among E-sepsis (**Figure 2**).

**Figure 2.**
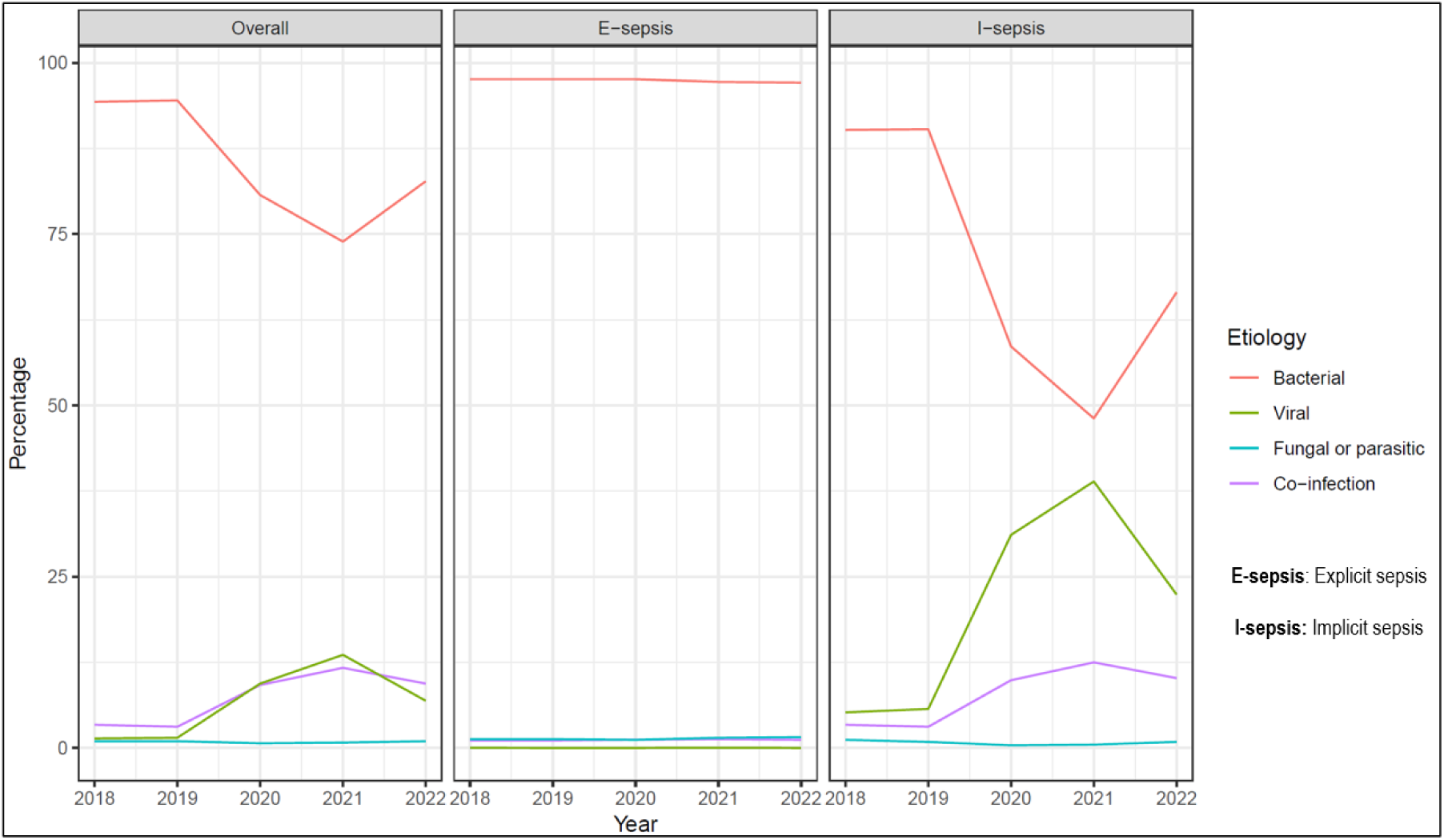
Distribution of sepsis etiology, overall and by type of sepsis, in metropolitan France, 2018-2022.

### Costs of sepsis-associated hospital stays

Total sepsis-related hospitalization costs and extra medication costs increased during the pandemic period (**Figure 3**). While the total costs for hospitalization and extra medication were higher for E-sepsis than I-sepsis (**Figure 3**), the median cost per hospital stay was higher for I-sepsis than E-sepsis (in 2022, median (IQR) cost was €13,094 (€7,670-€23,316) for I-sepsis and €8,233 (€4,898-€15,874) for E-sepsis) (**Figure 4** and **eTable 6**). The higher total costs during the pandemic period was mainly attributable to the increased total costs for I-sepsis.

**Figure 3.**
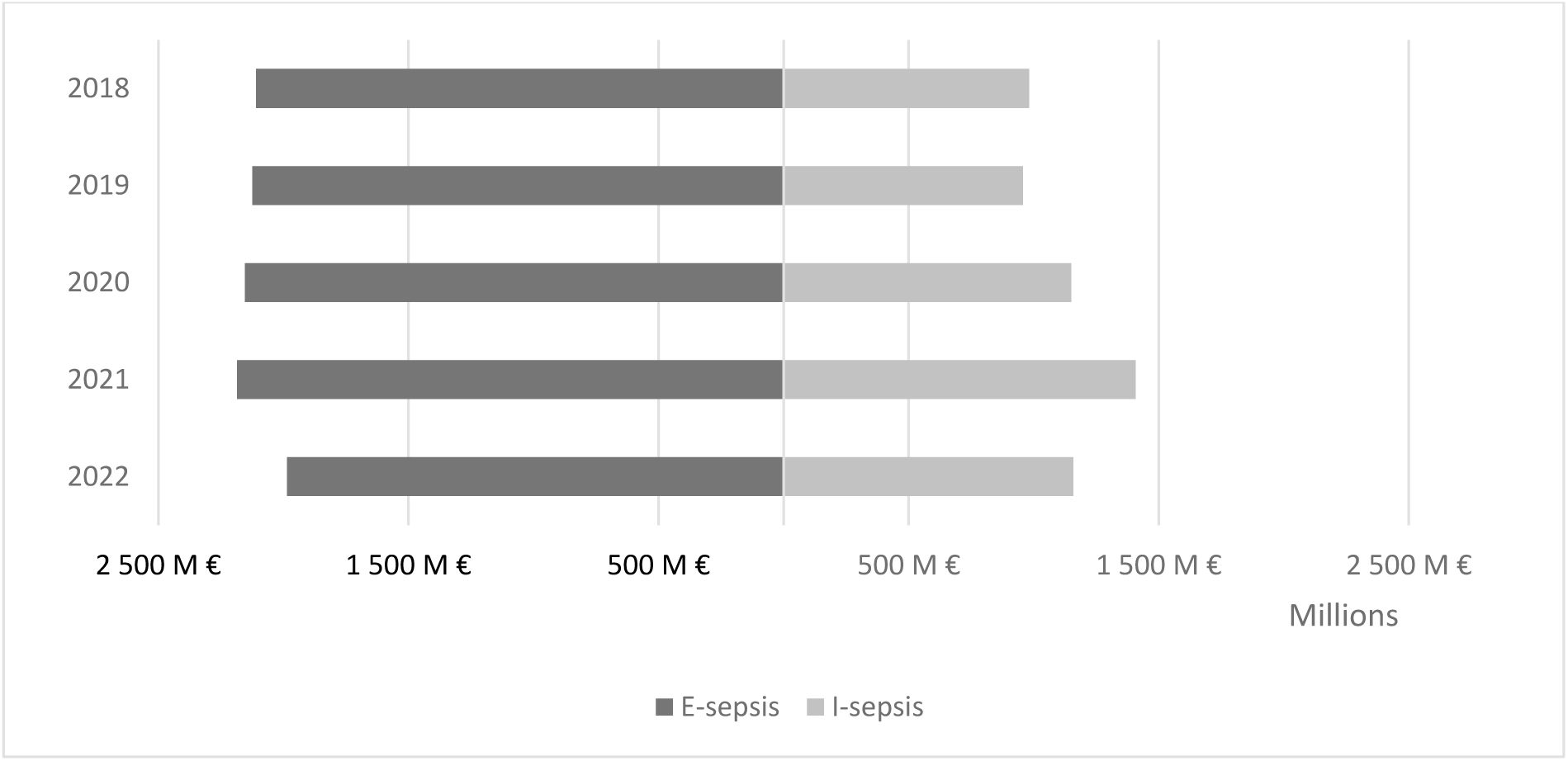
Total monetary cost of sepsis-associated hospitalization by sepsis type in metropolitan France, 2018-2022.

**Figure 4.**
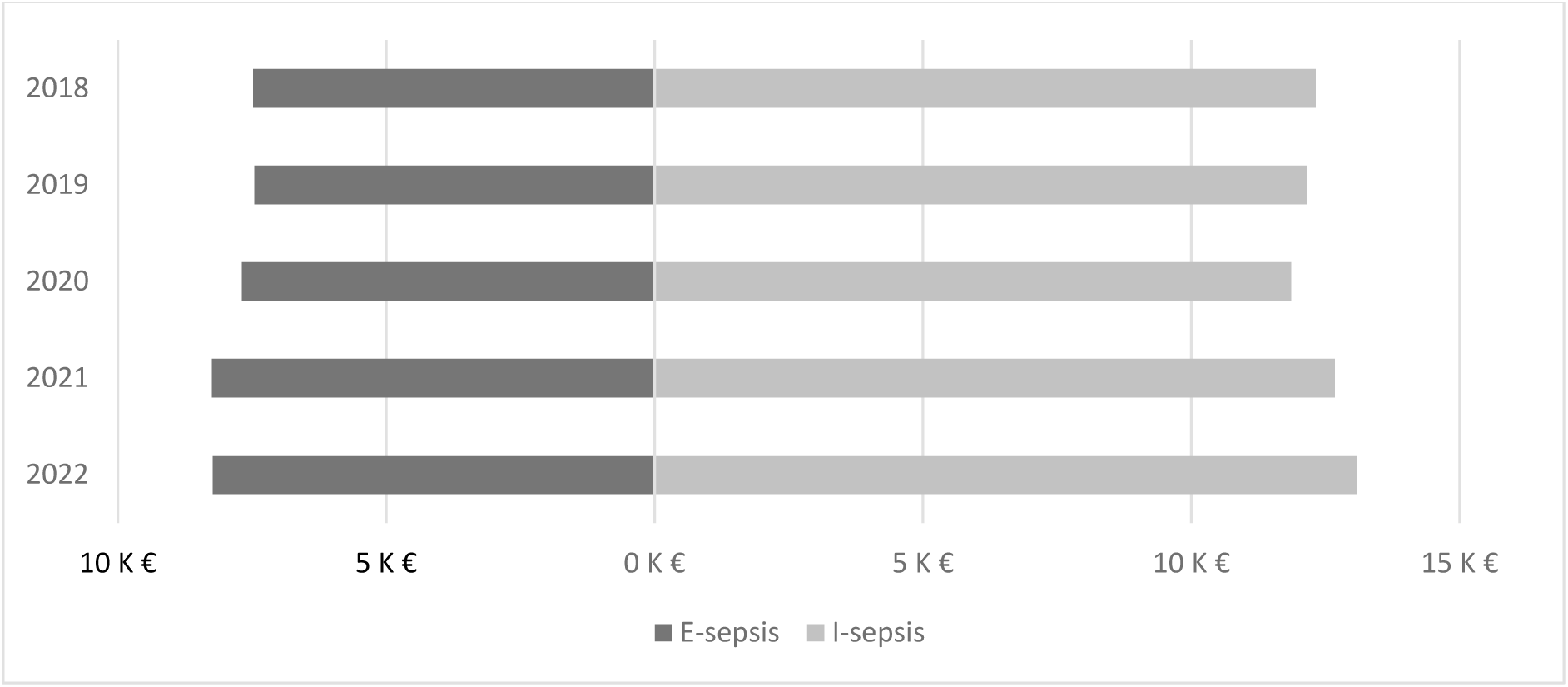
Median monetary cost of sepsis-associated hospitalization by sepsis type in metropolitan France, 2018-2022.

### Patient characteristics

Overall, patient characteristics over the five-year study period were stable apart from a marked increase in lower respiratory tract infections during the pandemic (**eTable 3**). Over the 5 years, in both type of sepsis patients, about two-thirds were aged between 55-85 years, with a consistently higher proportion of males than females. The most frequent comorbidities recorded were cancer, chronic heart failure, renal disease and chronic pulmonary disease, and the most frequent sites of infection were lower respiratory tract, urinary and genital tract and multiple sites (**eTable 3**).

Changes in patient characteristics before and during the pandemic were only noticeable in I-sepsis (**Table 1**). During the pandemic, among I-sepsis, there were more males, patients aged 55-85 years, patients having a Charlson index of 0 and patients with lower respiratory tract infection, while there were fewer patients with cancer, congestive heart failure, renal disease and chronic pulmonary disease and patients with multiple infection sites, urinary and genital tract and unknown site of infection, compared to the pre-pandemic period, although these trends began to reverse in 2022.

At least two doses of COVID-19 vaccine had been administered to 59% and 83% of sepsis patients by the end of 2021 and by the end of 2022, respectively (**eTable 5**).

### Hospital stay characteristics

The median length of stay was stable over the years for both E-sepsis and I-sepsis cases (around 12 and 14 days for E- and I-sepsis respectively) (**eTable 4**). More than 80% of patients were admitted from home for both types, with a slight decrease for I-sepsis during the pandemic (from 85.1% in 2018 to 80.4% in 2021), while patients coming from acute care increased (from 13.4% in 2018 to 18.4% in 2021). Considering E-sepsis only, the share of patient experiencing septic shock and ICU admission remained stable over the years.

Almost half of cases returned home after their hospitalization while almost a quarter died (**eTable 3**). The percentage of sepsis cases returning home slightly decreased during the COVID-19 pandemic whereas in-hospital mortality slightly increased (p <.0001). In-hospital mortality was consistently higher among E-sepsis than I-sepsis cases, but increased for both types during the pandemic period (from 18% and 25.2% in 2019 to 20% and 30.9% in 2022, respectively, for I- and E-sepsis; p <.0001 for both) (**eTable 4**). Patients with I-sepsis were more frequently transferred to acute or long-term care after hospitalization than those with E-sepsis.

## Discussion

Our study has estimated the burden of sepsis before and during the COVID-19 pandemic in metropolitan France using a national medico-administrative database. While before the pandemic sepsis incidence was observed to be stable with predominantly bacterial etiology, there was a modest increase in incidence in 2020 and 2021, mainly due to increased viral I-sepsis associated with SARS-CoV-2, representing more than 13% or more of all cases in 2021. Higher total sepsis-related hospitalization costs were also observed during the pandemic.

Before the pandemic, non-bacterial sepsis represented a minor percentage of all cases (2.5% in France, in our study, which is similar to the results of a previous study ^12^). However, rather than indicating the near absence of viral sepsis, extremely low rates of non-bacterial sepsis reported in most epidemiological studies likely reflects its lack of recognition, undertesting and diagnostic coding challenges. Since only two ICD-10 codes for viral-associated sepsis exist, and none for sepsis associated with SARS-CoV-2, the total incidence of viral-associated sepsis must be estimated by an indirect approach ^1,12^.

High rates of sepsis in COVID-19 patients have been widely reported previously. A systematic review and meta-analysis from early in the pandemic identified rates close to 30% of SARS-CoV-2-associated sepsis in general wards, reaching 78% in ICU patients ^17^. Despite this surge in viral sepsis, the increase in overall sepsis incidence that we observed during the pandemic was modest due to a significant concomitant drop in bacterial sepsis. This result is consistent with a nationwide study in England, which found a decrease in non-COVID-19 sepsis incidence during the pandemic followed by a rebound to pre-pandemic levels after lifting national lockdowns ^21^, as well as with surveillance data from 30 countries and territories (including France) participating in the Invasive Respiratory Infection Surveillance Consortium, which showed a sustained decrease in the incidence of invasive diseases caused by respiratory bacteria during the first two years of the pandemic ^22^. In 2022, our study showed a decrease in the number of sepsis cases due to a decrease in both bacterial and viral sepsis compared to previous years. While high COVID-19 vaccination coverage in France by 2022 likely contributed to this reduction in viral sepsis, the marked overall decrease from 2021 to 2022 may also be due to changes in coding practices in France following recommendations issued in 2021 (**eMethods**), in addition to some lasting infection prevention due to pandemic restrictions and behavior change. A recent English study showed that, even before pandemic, there was an impact of changes in sepsis coding guidance on reported trends, resulting in a small reduction in sepsis mortality rates across England ^23^.

In a previous review of 18 studies, overall per-patient sepsis hospitalization costs were estimated to be high, but highly variable between countries ^3^. The majority of studies were performed in the United States of America (USA) and in Europe. The median (IQR) of the total sepsis hospitalization costs per stay was €36,191 (€17,158-€53,349). A similar same range was estimated in our study, although an important difference between E-sepsis and I-sepsis cases was observed, with higher median costs for I-sepsis than E-sepsis, which could be related to its definition that includes ICU admission and organ dysfunction/support. In addition, total costs of sepsis hospital stays including extra medication costs increased during the pandemic, with a particularly notable increase for I-sepsis. That finding is in accordance with a study ^24^, which found that during the pandemic, COVID-19 patients with sepsis or pneumonia had higher costs than those without COVID-19.

Changes in the characteristics of patients and their stays during the pandemic were more significant for I-sepsis than E-sepsis. Compared with the pre-pandemic period, I-sepsis patients were more often males aged between 55-85 years with Charlson index of 0, and had fewer comorbidities (especially cancer, congestive heart failure, renal disease and chronic pulmonary disease) and more lower respiratory tract infection. These changes are likely related to SARS-CoV-2-associated sepsis, of which the profile might differ from that of the typical bacterial sepsis ^25^.

## Limitations of the study

Although we used a methodology of identifying sepsis in medico-administrative databases based on explicit and implicit diagnostic codes, as considered in previous studies ^1,12^, there are a number of limitations using this method, particularly the risk of misclassification for implicit sepsis. Also, the definition of sepsis case varied through the years, with new recommendations and guidelines for coding practices occurring intermittently ^6^. This can limit comparison between studies on sepsis and may explain at least in part the large heterogeneity in sepsis prevalence that the meta-analysis of Karakike et al ^17^ observed.

While the use of medico-administrative databases to analyze changes in sepsis incidence and mortality is debated ^26–28^, this approach remains the most cost-effective way to study sepsis at the national level. Our results suggesting an impact of the COVID-19 pandemic on the burden of sepsis is in agreement with findings of a recent study that used validated electronic clinical data from 5 Massachusetts hospitals to show that SARS-CoV-2 had a significant contribution to the burden of sepsis in hospitalized patients ^29^.

## Conclusion

During the COVID-19 pandemic, the overall incidence and economic burden of sepsis increased in French hospitals despite a significant reduction in bacterial sepsis. We identified viral sepsis using implicit diagnostic codes in a national electronic health records database. Although this methodology has been previously validated and is used, our study nonetheless highlights the lack of explicit codes for viral sepsis, which poses challenges for epidemiological analysis and could lead to underestimation of sepsis burden, particularly during pandemics or outbreaks when diagnostics and pathogen-specific codes may be unavailable.

## Data Availability

The data that support the findings of this study are available from the French administrative health care database (SNDS) but restrictions apply to the availability of these data, which were used under the French Data Protection Agency approval, and so are not publicly available.

## Authors Contributions

*Concept and design: Watier, Brun-Buisson*.

*Acquisition, analysis, or interpretation of data: Al Rahmoun, Watier, Brun-Buisson.*

*Drafting of the manuscript: Al Rahmoun, Watier, Brun-Buisson*.

*Critical review of the manuscript for important intellectual content: Elabbadi, Guillemot, Watier, Brun-Buisson*.

*Statistical analysis: Al Rahmoun,*

*Obtained funding: Watier*.

*Administrative, technical, or material support: Guillemot, Watier.*

*Supervision: Watier*.

## Acknowledgements/ Additional Contributions

We are grateful to the DATAD department of the French National Health Insurance for providing the data and to David R. M. Smith for his proofreading of the manuscript.

