## Supplementary data for "Impacts of the COVID-19 pandemic on sepsis incidence, etiology and hospitalization costs in France: a retrospective observational study"

### eMethods

#### Description of the French National Hospital Discharge Database (PMSI)

This study was approved by the French Data Protection Agency (CNIL, approval DR-2022-146). Informed consent is waived for the use of these anonymized secondary data, as mentioned in the Social Security Code, Article L161-28-1. All methods were performed in accordance with CNIL regulations and with REporting of studies Conducted using Observational Routinely collected Data (RECORD) guidelines. The SNDS (Système national des données de santé), a national medico-administrative database, contains individual data used for billing and reimbursement of outpatient health care consumption (Inter-Scheme consumption data: DCIR) and private and public hospital data (Medical Information System Program: PMSI) collected by the Agence technique de l'information sur l'hospitalisation (ATIH)[1].

PMSI data include hospital discharge summaries covering all hospital stays in publicly funded and private institutions, including acute-care facilities (medicine, surgery or obstetrics units: MSO)[1]. For each stay, the diagnoses are coded with ICD-10-codes as primary diagnosis (PD: condition requiring hospitalization), related diagnosis (RD: adds information to PD) and significant associated diagnosis (SAD: complications and co-morbidities potentially affecting the course or cost of hospitalization). While PD and RD are unique for each stay, several SAD can be attributed per stay. Additional information is available about the patients, such as sex or age and about the hospital stays as entry and exit date, admission source, hospital discharge (including death) or medical procedures. Recommendations about coding practices are regularly published by the ATIH. Recommendations on coding practices for sepsis were published in 2014, especially concerning the use of R65.1 and R57.2 ICD-10 codes combined with infection codes in order to better identify organ dysfunction and septic shock [2]. Further recommendations about coding practices for sepsis were updated in 2021 [3]. These updated recommendations, following the application of the 2016 definition of sepsis, resulted in eliminating codes R650 or R651 from the sepsis definition and were applied in the current study.

**eTable 1. ICD-10 codes used to identify sepsis of presumed bacterial, viral, fungal or parasitic etiology according to type of selection**

| **E-sepsis codes** [a,b,c,e] | **I-sepsis** [a,c,d,e] | | |
|---|---|---|---|
| | **Infection codes** [b] | **1st associated condition** | **2nd associated condition** |
| **Sepsis of presumed bacterial etiology** | | | |
| A021, A227, A267, A327, A400-A409, A410-A419, A427, A480, A483, O85, O883, R572, R578 | A040-A049, A390-A399, G000-G009, I330, J068, J13, J14, J150-J159, J160-J168, J180-J189, J869, K650, K659, K810, K830, L022, L089, M000-M0099, M4620-M4629, M6000-M6009, M8600-M8609, M8690-M8699, N136, N390, T793, T802, T811, T814, T827, T845, T857 | ICU admission | ICD-10 codes for organ dysfunction: D65, D689, D695, D696, D762, E86, E872, F05, F09, G934, I460, I469, I959, J80, J81, J952, J9600, J9601, J9609, K720, K729, N080, N088, N160, N17, N19, R092, R17, R34, R40, R392, R410, R418, R55, R571, R579<br><br>CCAM codes for organ support: DKMD001, DKMD002, EQLF002, EQLF003, FELF003, GLLD003, GLLD004, GLLD008, GLLD012, GLLD015, GLLD019, JVJB002, JVJF002, JVJF003, JVJF005 |
| **Sepsis of presumed viral etiology** | | | |
| A99, B007 | A86, A87, A91, A92, A94, A96, A98, B01, B25, B27, J09, J10, J11, J12, B334, B341, U049, U109[f], U0710[f], U0711[f] | *same* | *same* |
| **Sepsis of presumed fungal or parasitic etiology** | | | |
| B377, B508 | B45, B49, B58, B59, B387, B393, B407, B440, B441, B447, B448, B449, B457, B464, B487, B787, B509, B500 | *same* | *same* |

[a] E-sepsis: Explicit sepsis, I-sepsis: Implicit sepsis.

[b] One of the ICD-10 code as primary diagnosis (PD: condition requiring hospitalization), related diagnosis (RD: adds information to PD) or significant associated diagnosis (SAD: complications and co-morbidities potentially affecting the course or cost of hospitalization).

[c] Sepsis = explicit sepsis + implicit sepsis.

[d] Implicit sepsis = ICD-10 code of infection + ICU admission + organ dysfunction/support.

[e] Stays shorter than 24h hours without death were excluded from our selection.

[f] Codes for Covid-19 infection.

**eTable 2. Yearly number of hospital stays (reported as % of sepsis cases) for sepsis patients hospitalized in metropolitan France, 2018-2022**

| | 2018 | 2019 | 2020 | 2021 | 2022 |
|---|---|---|---|---|---|
| **Number of stays** | | | | | |
| 1 | 88.2% | 87.8% | 87.7% | 89.4% | 89.8% |
| 2 | 9.5% | 9.8% | 9.9% | 8.9% | 8.5% |
| >2 | 2.3% | 2.4% | 2.4% | 1.7% | 1.7% |

**eTable 3. Characteristics of all patients and hospital stays with sepsis, metropolitan France 2018-2022**

| | **2018 (n=229,668)** | **2019 (n=232,355)** | **2020 (n=242,175)** | **2021 (n=236,076)** | **2022 (n=207,409)** |
|---|---|---|---|---|---|
| ***Patient's characteristics, n (%)*** | | | | | |
| **Sex** | | | | | |
| Male | 132,634 (57.8) | 134,393 (57.8) | 145,487 (60.1) | 142,733 (60.5) | 123,952 (59.8) |
| Female | 97,034 (42.2) | 97,962 (42.2) | 96,688 (39.9) | 93,343 (39.5) | 83,457 (40.2) |
| **Age (years)** | | | | | |
| 15-29 | 6,733 (2.9) | 6,660 (2.9) | 6,065 (2.5) | 6,025 (2.6) | 5,662 (2.7) |
| 30-44 | 12,655 (5.5) | 12,305 (5.3) | 12,692 (5.2) | 13,960 (5.9) | 11,101 (5.4) |
| 45-54 | 18,616 (8.1) | 18,403 (7.9) | 20,015 (8.3) | 21,023 (8.9) | 15,712 (7.6) |
| 55-64 | 37,405 (16.3) | 36,963 (15.9) | 40,274 (16.6) | 40,895 (17.3) | 32,942 (15.9) |
| 65-74 | 56,831 (24.8) | 58,653 (25.2) | 64,846 (26.8) | 65,275 (27.7) | 55,946 (27.0) |
| 75-84 | 55,169 (24.0) | 55,471 (23.9) | 56,757 (23.4) | 52,230 (22.1) | 49,863 (24.0) |
| ≥ 85 | 42,259 (18.4) | 43,900 (18.9) | 41,526 (17.2) | 36,668 (15.5) | 36,183 (17.4) |
| Median [IQR] | 71 [60-82] | 72 [61-82] | 71 [61-81] | 70 [60-80] | 72 [61-81] |
| **Charlson index** | | | | | |
| 0 | 78,016 (34.0) | 79,069 (34.0) | 87,971 (36.3) | 91,391 (38.7) | 68,824 (33.2) |
| 1-2 | 82,087 (35.7) | 83,294 (35.9) | 85,217 (35.2) | 80,231 (34.0) | 74,978 (36.2) |
| 3-4 | 34,185 (14.9) | 34,477 (14.8) | 34,569 (14.3) | 33,014 (14.0) | 33,201 (16.0) |
| ≥ 5 | 35,380 (15.4) | 35,515 (15.3) | 34,418 (14.2) | 31,440 (13.3) | 30,406 (14.7) |
| Median [IQR] | 2 [0-3] | 2 [0-3] | 2 [0-3] | 2 [0-3] | 2 [0-3] |
| **Comorbidities** | | | | | |
| Cancer | 56,189 (24.5) | 56,302 (24.2) | 54,470 (22.5) | 49,184 (20.8) | 47,177 (22.8) |
| Congestive heart failure | 49,777 (21.7) | 49,857 (21.5) | 50,224 (20.7) | 48,874 (20.7) | 48,883 (23.6) |
| Renal disease | 30,599 (13.3) | 31,718 (13.7) | 32,619 (13.5) | 31,675 (13.4) | 31,341 (15.1) |
| Chronic pulmonary disease | 27,097 (11.8) | 27,545 (11.9) | 28,358 (11.7) | 27,966 (11.9) | 28,206 (13.6) |
| Metastatic carcinoma | 23,378 (10.2) | 23,676 (10.2) | 22,850 (9.4) | 20,340 (8.6) | 19,114 (9.2) |
| Diabetes with chronic complications | 13,772 (6.0) | 13,825 (5.9) | 14,220 (5.9) | 13,270 (5.6) | 11,925 (5.8) |
| Paraplegia or hemiplegia | 13,270 (5.8) | 13,311 (5.7) | 13,808 (5.7) | 13,343 (5.7) | 12,161 (5.9) |
| Dementia | 11,865 (5.2) | 11,814 (5.1) | 11,305 (4.7) | 9,691 (4.1) | 9,570 (4.6) |
| Mild liver disease | 12,426 (5.4) | 12,779 (5.5) | 13,010 (5.4) | 12,396 (5.3) | 11,863 (5.7) |
| Moderate or severe liver disease | 5,900 (2.6) | 5,839 (2.5) | 5,757 (2.4) | 5,320 (2.3) | 5,494 (2.7) |
| Rheumatological disease | 2,859 (1.2) | 2,943 (1.3) | 3,045 (1.3) | 2,983 (1.3) | 2,775 (1.3) |
| | | | | | *(continued)* |
| AIDS | 1,113 (0.5) | 1,033 (0.4) | 1,028 (0.4) | 895 (0.4) | 830 (0.4) |

**eTable 3. Characteristics of all patients and hospital stays with sepsis, metropolitan France 2018-2022**

| | 2018 (n=229,668) | 2019 (n=232,355) | 2020 (n=242,175) | 2021 (n=236,076) | 2022 (n=207,409) |
|---|---|---|---|---|---|
| **Infection sites** | | | | | |
| Multiple sites | 51,079 (22.2) | 51,718 (22.3) | 49,820 (20.6) | 46,671 (19.8) | 46,760 (22.5) |
| Lower respiratory tract | 46,192 (20.1) | 45,727 (19.7) | 70,773 (29.2) | 82,166 (34.8) | 58,153 (28.0) |
| Blood | 36,533 (15.9) | 36,634 (15.8) | 33,001 (13.6) | 29,611 (12.5) | 27,796 (13.4) |
| Urinary and genital tract | 34,945 (15.2) | 35,904 (15.5) | 33,422 (13.8) | 28,972 (12.3) | 26,743 (12.9) |
| Gastrointestinal and abdomen | 14,232 (6.2) | 14,516 (6.3) | 13,683 (5.7) | 12,296 (5.2) | 11,280 (5.4) |
| Heart and mediastinum | 11,248 (4.9) | 11,546 (5.0) | 10,592 (4.4) | 8,803 (3.7) | 8,062 (3.9) |
| Skin and soft tissues | 10,940 (4.8) | 11,205 (4.8) | 9,923 (4.1) | 9,010 (3.8) | 8,743 (4.2) |
| Medical devices | 7,865 (3.4) | 7,909 (3.4) | 7,073 (2.9) | 6,171 (2.6) | 5,409 (2.6) |
| Bones and joints | 4,716 (2.1) | 5,005 (2.2) | 4,401 (1.8) | 4,041 (1.7) | 3,925 (1.9) |
| Nervous system | 897 (0.4) | 931 (0.4) | 652 (0.3) | 605 (0.3) | 713 (0.3) |
| Ears, nose and throat | 364 (0.2) | 365 (0.2) | 244 (0.1) | 219 (0.1) | 223 (0.1) |
| Pregnancy | 314 (0.1) | 314 (0.1) | 283 (0.1) | 269 (0.1) | 266 (0.1) |
| Eyes | 34 (0.0) | 30 (0.0) | 24 (0.0) | 28 (0.0) | 23 (0.0) |
| Unknown | 10,309 (4.5) | 10,551 (4.5) | 8,284 (3.4) | 7,214 (3.1) | 9,313 (4.5) |
| ***Hospital stay's characteristics, n (%)*** | | | | | |
| **Admission source** | | | | | |
| Home | 196,327 (85.5) | 198,541 (85.5) | 203,739 (84.2) | 195,690 (82.9) | 174,809 (84.3) |
| Acute care | 28,229 (12.3) | 28,721 (12.3) | 33,501 (13.8) | 36,036 (15.3) | 28,638 (13.8) |
| Long-term care | 5,112 (2.2) | 5,093 (2.2) | 4,935 (2.0) | 4,350 (1.8) | 3,962 (1.9) |
| **Length of stay (days)** | | | | | |
| <7 | 54,997 (24.0) | 56,279 (24.2) | 56,313 (23.3) | 53,128 (22.5) | 50,686 (24.4) |
| 7-14 | 70,992 (30.9) | 72,115 (31.0) | 76,096 (31.4) | 75,278 (31.9) | 62,778 (30.3) |
| 15-30 | 65,753 (28.6) | 66,128 (28.5) | 70,818 (29.2) | 68,004 (28.8) | 58,161 (28.0) |
| >30 | 37,926 (16.5) | 37,833 (16.3) | 38,948 (16.1) | 39,666 (16.8) | 35,784 (17.3) |
| Median [IQR] | 13 [7-24] | 13 [7-23] | 13 [7-23] | 13 [7-24] | 13 [7-24] |
| **ICU admission[a]** | | | | | |
| Yes | 130,948 (57.0) | 130,540 (56.2) | 144,505 (59.7) | 149,986 (63.5) | 128,587 (62.0) |
| No | 98,720 (43.0) | 101,815 (43.8) | 97,670 (40.3) | 86,090 (36.5) | 78,822 (38.0) |
| **Septic shock[a]** | | | | | |
| Yes | 52,690 (22.9) | 53,328 (23..0) | 52,467 (21.7) | 49,579 (21.0) | 49,365 (23.8) |
| No | 176,978 (77.1) | 179,027 (77.0) | 189,708 (78.3) | 186.497 (79.0) | 158,044 (76.2) |
| **Hospital discharge** | | | | | |
| Home | 111,786 (48.7) | 114,054 (49.1) | 116,081 (47.9) | 111,303 (47.2) | 96,273 (46.4) |
| Acute care | 27,746 (12.1) | 27,267 (11.7) | 26,633 (11.0) | 25,900 (11.0) | 22,818 (11.0) |
| Long-term care | 35,903 (15.6) | 36,625 (15.8) | 39,583 (16.3) | 37,252 (15.8) | 31,115 (15.0) |
| Death | 54,233 (23.6) | 54,409 (23.4) | 59,878 (24.7) | 61,621 (26.1) | 57,203 (27.6) |

Abbreviation: IQR, interquartile range.

[a] Only among E-sepsis.

**eTable 4. Characteristics of hospital stays with sepsis according to sepsis type (E-sepsis/I-sepsis)[a] in metropolitan France, 2018-2022**

| | **2018 (n=229,668)** | | **2019 (n=232,355)** | | **2020 (n=242,175)** | | **2021 (n=236,076)** | | **2022 (n=207,409)** | |
|---|---|---|---|---|---|---|---|---|---|---|
| | E-sepsis (n=170,480) | I-sepsis (n=59,188) | E-sepsis (n=174,062) | I-sepsis (n=58,293) | E-sepsis (n=168,953) | I-sepsis (n=73,222) | E-sepsis (n=153,784) | I-sepsis (n=82,292) | E-sepsis (n=144,036) | I-sepsis (n=63,373) |
| **Admission source** | | | | | | | | | | |
| Home | 145,982 (85.6) | 50,345 (85.1) | 148,900 (85.5) | 49,641 (85.2) | 143,568 (85.0) | 60,171 (82.2) | 129,536 (84.2) | 66,154 (80.4) | 122,687 (85.2) | 52,122 (82.3) |
| Acute care | 20,283 (11.9) | 7,946 (13.4) | 20,973 (12.1) | 7,748 (13.3) | 21,505 (12.7) | 11,996 (16.4) | 20,859 (13.6) | 15,177 (18.4) | 18,187 (12.6) | 10,451 (16.5) |
| Long-term care | 4,215 (2.5) | 897 (1.5) | 4,189 (2.4) | 904 (1.6) | 3,880 (2.3) | 1,055 (1.4) | 3,389 (2.2) | 961 (1.2) | 3,162 (2.2) | 800 (1.3) |
| **Length of stay (days)** | | | | | | | | | | |
| <7 | 45,700 (26.8) | 9,297 (15.7) | 46,841 (26.9) | 9,438 (16.2) | 44,086 (26.1) | 12,227 (16.7) | 40,185 (26.1) | 12,943 (15.7) | 39,646 (27.5) | 11,040 (17.4) |
| 7-14 | 51,762 (30.4) | 19,230 (32.5) | 53,107 (30.5) | 19,008 (32.6) | 50,967 (30.1) | 25,129 (34.3) | 45,474 (29.6) | 29,804 (36.2) | 42,121 (29.2) | 20,657 (32.6) |
| 15-30 | 45,867 (26.9) | 19,886 (33.6) | 46,673 (26.8) | 19,455 (33.4) | 46,461 (27.5) | 24,357 (33.3) | 41,677 (27.1) | 26,327 (32.0) | 38,222 (26.5) | 19,939 (31.5) |
| >30 | 27,151 (15.9) | 10,775 (18.2) | 27,441 (15.8) | 10,392 (17.8) | 27,439 (16.2) | 11,509 (15.7) | 26,448 (17.2) | 13,218 (16.1) | 24,047 (16.7) | 11,737 (18.5) |
| Median (IQR) | 12 [6-23] | 15 [9-25] | 12 [6-23] | 15 [9-25] | 13 [6-23] | 14 [8-24] | 13 [6-24] | 14 [9-24] | 12 [6-23] | 14 [8-25] |
| **ICU admission[b]** | | | | | | | | | | |
| Yes | 71,760 (42.1) | 59,188 (100) | 72,247 (41.5) | 58,293 (100) | 71,283 (42.2) | 73,222 (100) | 67,694 (44.0) | 82,292 (100) | 65,214 (45.3) | 63,373 (100) |
| No | 98,720 (57.9) | - | 101,815 (58.5) | - | 97,670 (57.8) | - | 86,090 (56.0) | - | 78,822 (54.7) | - |
| **Septic shock[b]** | | | | | | | | | | |
| Yes | 52,690 (30.9) | - | 53,328 (30.6) | - | 52,467 (31.0) | - | 49,579 (32.2) | - | 49,365 (34.3) | - |
| No | 117,790 (69.1) | 59,188 (100) | 120,734 (69.4) | 58,293 (100) | 116,486 (69.0) | 73,222 (100) | 104,205 (67.8) | 82,292 (100) | 94,671 (65.7) | 63,373 (100) |
| **Hospital discharge** | | | | | | | | | | |
| Home | 85,113 (49.9) | 26,673 (45.1) | 87,451 (50.2) | 26,603 (45.6) | 82,624 (48.9) | 33,457 (45.7) | 71,324 (46.4) | 39,979 (48.6) | 66,306 (46.0) | 29,967 (47.3) |
| Acute care | 16,711 (9.8) | 11,035 (18.6) | 16,960 (9.7) | 10,307 (17.7) | 15,359 (9.1) | 11,274 (15.4) | 14,103 (9.2) | 11,797 (14.3) | 13,156 (9.1) | 9,662 (15.3) |
| Long-term care | 87,169 (14.6) | 11,077 (18.7) | 25,751 (14.8) | 10,874 (18.7) | 25,883 (15.3) | 13,700 (18.7) | 22,890 (14.9) | 14,362 (17.5) | 20,087 (13.9) | 11,028 (17.4) |
| Death | 43,830 (25.7) | 10,403 (17.6) | 43,900 (25.2) | 10,509 (18.0) | 45,087 (26.7) | 14,791 (20.2) | 45,467 (29.6) | 16,154 (19.6) | 44,487 (30.9) | 12,716 (20.1) |

Abbreviations: IQR, interquartile range ; ICU, intensive care unit.
[a] E-sepsis: Explicit sepsis, I-sepsis: Implicit sepsis.[b] Only in E-sepsis.

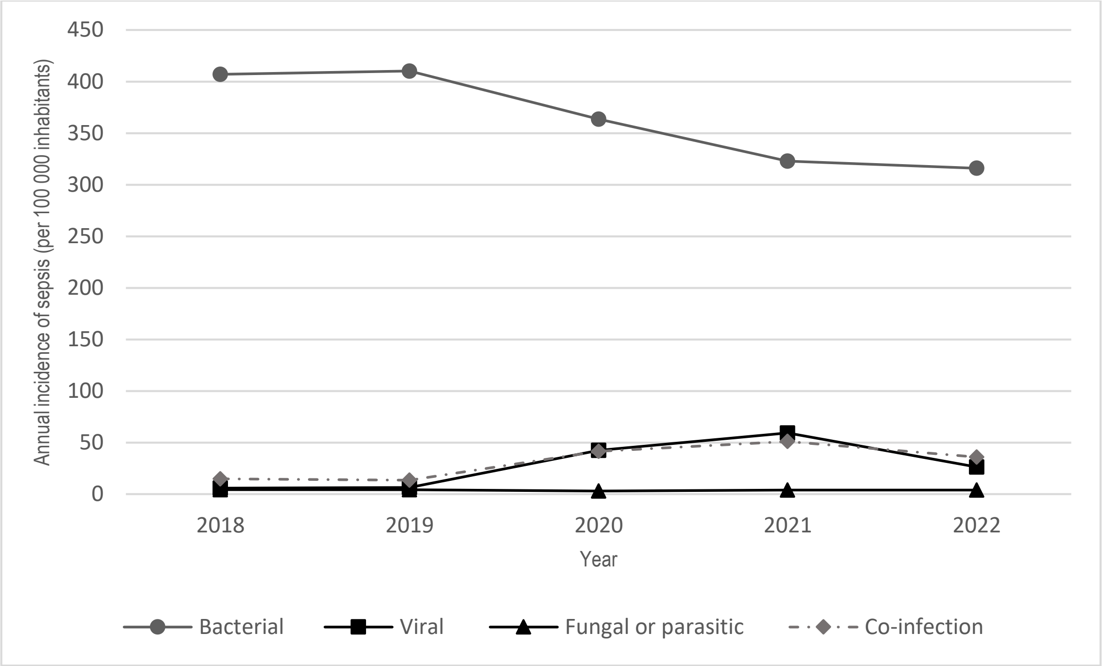


**eFigure 1. Sepsis incidence rate according to etiology in metropolitan France, 2018-2022**

**eTable 5. Covid-19 vaccine coverage among cases of sepsis in 2021 and 2022**

| Number of Covid-19 vaccine doses | 2021 (n=236,076) | | 2022 (n=207,409) | |
|---|---|---|---|---|
| 0 | 66,814 | 28.3 | 29,527 | 14.2 |
| 1 | 31,262 | 13.2 | 5,334 | 2.6 |
| 2 | 77,116 | 32.7 | 27,569 | 13.3 |
| 3 | 59,334 | 25.1 | 93,159 | 44.9 |
| ≥ 4 | 1,550 | 0.7 | 51,820 | 25.0 |

**eTable 6. Cost of hospital stays with sepsis**

| Hospital stay cost (euros) | E-sepsis | I-sepsis | Total |
|---|---|---|---|
| **2018** | | | |
| Cost per stay [median, IQR] | 7,479 [4,521-14,409] | 12,315 [6,877-21,331] | 8,436 [5,076-16,446] |
| Total cost without extra medication | 2,108,997,759 | 981,202,385 | 3,090,200,144 |
| Total extra medication costs | 58,001,989 | 15,137,662 | 73,139,650 |
| Total cost with extra medication | 2,166,999,748 | 996,340,047 | 3,163,339,794 |
| **2019** | | | |
| Cost per stay [median, IQR] | 7,454 [4,519-14,127] | 12,151 [6,899-21,027] | 8,342 [5,075-16,111] |
| Total cost without extra medication | 2,123,677,196 | 957,265,632 | 3,080,942,827 |
| Total extra medication costs | 68,240,960 | 19,159,512 | 87,400,471 |
| Total cost with extra medication | 2,191,918,156 | 976,425,144 | 3,168,343,298 |
| **2020** | | | |
| Cost per stay [median, IQR] | 7,688 [4,650-14,978] | 11,857 [7,396-19,982] | 8,788 [5,439-16,719] |
| Total cost without extra medication | 2,154,882,126 | 1,150,283,326 | 3,305,165,452 |
| Total extra medication costs | 77,248,881 | 14,666,765 | 91,915,646 |
| Total cost with extra medication | 2,232,131,007 | 1,164,950,091 | 3,397,081,098 |
| **2021** | | | |
| Cost per stay [median, IQR] | 8,252 [4,919-16,380] | 12,673 [8,021-21,126] | 9,749 [6,028-18,322] |
| Total cost without extra medication | 2,185,018,233 | 1,407,864,954 | 3,592,883,186 |
| Total extra medication costs | 75,463,226 | 25,711,983 | 101,175,209 |
| Total cost with extra medication | 2,260,481,459 | 1,433,576,937 | 3,694,058,395 |
| **2022** | | | |
| Cost per stay [median, IQR] | 8,233 [4,898-15,874] | 13,094 [7,670-23,316] | 9,431 [5,687-18,356] |
| Total cost without extra medication | 1,985,392,191 | 1,157,733,418 | 3,143,125,609 |
| Total extra medication costs | 42,058,833 | 21,114,791 | 63,173,624 |
| Total cost with extra medication | 2,027,451,024 | 1,178,848,209 | 3,206,299,233 |

Abbreviation: IQR, interquartile range.
